# Quantifying evidence for phenotypic specificity (PP4) for syndromic phenotypes: Large-scale integration of rare germline *FH* variants from diagnostic laboratory testing for HLRCC (Hereditary Leiomyomatosis and Renal Cell Cancer) and renal cancer

**DOI:** 10.1101/2025.03.17.25324088

**Authors:** Sophie Allen, Charlie F. Rowlands, Samantha Butler, Miranda Durkie, Carrie Horton, Tina Pesaran, Marcy Richardson, Rachel Robinson, Alice Garrett, George J. Burghel, Alison Callaway, Joanne Field, Bethan Frugtniet, Sheila Palmer-Smith, Jonathan Grant, Judith Pagan, Trudi McDevitt, Katie Snape, Avgi Andreaou, Eamonn R. Maher, Helen Hanson, Terri McVeigh, Clare Turnbull, CanVIG-UK

## Abstract

**Purpose:** Hereditary Leiomyomatosis and Renal Cell Cancer (HLRCC) is a rare cancer susceptibility syndrome exclusively attributable to pathogenic variants in *FH*. This paper quantitatively weights the phenotypic context (PP4/PS4) of such very rare variants in *FH*.

**Methods:** We collated clinical diagnostic testing data on germline *FH* variants from 387 individuals with HLRCC and 1,780 individuals with renal cancer, and compared the frequency of ‘very rare’ variants in each phenotypic cohort against 562,295 population controls. We generated pan-gene very rare variant likelihood ratios (PG-VRV-LRs), domain-specific likelihood ratios for missense variants (DS-VRMV-LR) using spatial clustering analysis, and log_2.08_ likelihood ratios (LLRs) as applicable within the updated ACMG/AMP Variant Classification Framework.

**Results:** For HLRCC, the PG-VRV-LR was estimated to be 2,669.4 (95% CI: 1,843.4-3,881.2, LLR 10.77) for truncating variants and 214.7 (185.0-246.9, LLR 7.33) for missense variants. For renal cancer, the PG-VRV-LR was 95.5 (48.9-183.0, LLR 6.23) for truncating variants and 5.8 (3.5-9.3, LLR 2.39) for missense variants. Clustering analysis in HLRCC cases revealed three ‘hotspot’ regions wherein the DS-VRMV-LR increased to 1226.9.

**Conclusion:** These data provide quantitative measures for very rare missense and truncating variants in *FH*, which reflect the differing phenotypic specificity of HLRCC and renal cancer, and may be applicable in clinical variant classification.

## Background

Hereditary Leiomyomatosis and Renal Cell Cancer (HLRCC) is a rare cancer susceptibility syndrome arising due to constitutional pathogenic variants in the *FH* (Fumarate hydratase) gene. The syndrome is characterised by cutaneous leiomyomas, uterine leiomyomas and susceptibility to renal cell cancers (most characteristically of type II papillary histology but with some other types described and recently updated WHO definition of pathological architecture)(1–3). Pheochromocytoma and paraganglioma have also been associated with *FH* pathogenic variants (PVs) in some cases. Germline biallelic *FH* variants cause an autosomal recessive neurometabolic disorder, fumarase deficiency (FMRD; MIM 606812).

*FH* is the only gene that has been associated with the HLRCC constellation of features. Hence, where the pattern of phenotypic features in an individual +/-their family are suggestive of HLRCC, genetic testing is typically focused on the *FH* gene. In many genomics services, phenotypic-based criteria have been defined to determine eligibility for testing of cancer susceptibility genes. For example, there are standardised criteria within the NHS England National Test Directory under eligibility code R365 (testing of the *FH* gene for HLRCC (fumarate hydratase-related tumour syndromes), see Box 1(4). Analysis of *FH* may also be performed as part of a substantially larger gene panel for genetic evaluation in an individual presenting with renal cancer.

Variants that are predicted to truncate the protein and result in nonsense-mediated decay (including nonsense, canonical splicing and out-of-frame del-ins) characteristically result in loss of protein function and overall as a group arise highly infrequently in the general population. Variants of this mutational type are thus awarded high weighting towards a classification of (likely) Pathogenic in regard of this mechanism typically being strongly linked to the corresponding clinical disease. Conversely, missense variants are overall more likely to be neutral than deleterious and as a group more frequently observed in the general population. Mutational mechanism does not therefore typically advance assignation of a missense variant as pathogenic: evidence of other types is required.

That a given variant has been observed in an individual with the respective phenotype *can* be important evidence that the variant is disease-causing (or disease associated). The weighting assigned to this observation logically would be inherent to the specifics of the gene-phenotype-variant type triad, namely the rarity of the phenotype, the attribution of (deleterious variants in) that gene to the phenotype and the constraint of the gene for the given variant type. In the 2015 ACMG/AMP variant interpretation framework published by Richards et al, a default weighting of ‘supporting’ was assigned for when a “patient’s phenotype or family history is highly specific for a gene” (code PP4)(5). This weighting was acknowledged as deliberately conservative due to historic overweighting of phenotype information. In current practice, the degree to which PP4 (or variably PS4) is scored for phenotypic observations varies widely by disease with any standardised methodology. We have previously considered this question in the context of the very rare endocrine tumours phaeochromocytomas/paragangliomas (PHAEO-PGL) and the PHAEO-PGL-linked genes *SDHB* and *SDHD*. We analysed the frequency of very rare (i.e. plausibly pathogenic) missense variants in *SDHB* and *SDHD* in clinically-tested series of individuals with PHAEO-PGL and individuals in the general population, quantifying the phenotypic specificity for this exemplar scenario as a ‘very rare missense variant likelihood ratio’ (VRMV-LR). We sought to explore whether this methodology could be extended to other genes associated with more complex rare syndromic phenotypes.

Here, using data from four diagnostic laboratories, we examine the frequencies of rare missense and truncating *FH* variants in 387 cases of HLRCC, 1780 individuals with renal cancer undergoing clinical panel testing and 562,295 population controls. We used these data to explore the strength of evidence towards classification of pathogenicity that might be allocated for a rare missense variant in *FH* that is identified in (i) the high-specificity phenotypic context of HLRCC, comparing this to (ii) identification of a rare *FH* truncating variant in HLRCC and then comparing to identification of these *FH* variant types in (iii) the lower-specificity phenotype of renal cancer.

#### Box1: NHS National genomic test directory: Eligibility for R365: Testing for Fumarate hydratase-related tumour syndromes

The proband has:

a. Type 2 papillary OR tubulo-papillary renal tumour at any age, OR
b. Two of: cutaneous leiomyoma, renal tumour (any histology) OR uterine leiomyomas at any age, OR
c. Cutaneous leiomyoma AND one first / second / third degree relative with renal tumour, OR
d. Cutaneous leiomyoma AND two first / second / third degree relatives with cutaneous leiomyomas OR uterine leiomyomas with classic histological features < 40 years, OR
e. Uterine leiomyoma with classic histological features (age <40)
f. Multiple cutaneous leiomyomas

[Cutaneous leiomyomas should be histologically confirmed; uterine leiomyomas and renal tumours should be medically documented]

## Methods

### Assembly of clinical and laboratory expert group

The Cancer Variant Group UK (CanVIG-UK) is a group of >200 clinical diagnostic scientists and genetics clinicians working in the NHS in the UK and Republic of Ireland that has convened monthly since 2017(6). The CanVIG-UK Steering and Advisory Group (CStAG), which provides oversight for CanVIG-UK, comprises 13 senior clinical and laboratory experts providing representation from across all seven English Genomic Laboratory Hubs, as well as Scotland, Wales, and Ireland. The CStAG group, in combination with EM and AA (national clinical experts in HLRCC), provided input into the sourcing, analysis and interpretation of UK *FH* case data.

### Assembly of case data series

De-identified *FH* germline diagnostic genetic testing data was collated from three UK diagnostic laboratories (Birmingham, Sheffield, and Leeds) who perform the majority of *FH* testing in the UK, and one US-based central testing laboratory (Ambry) on the basis of (i) testing of *FH* alone on the basis of HLRCC-like phenotype or (ii) testing of *FH* within a Renal Cancer multigene panel test on the basis of renal cancer phenotype.

The Birmingham dataset comprised per-patient data from unrelated probands with renal cancer or suspected HLRCC referred to the West Midlands Regional Genomic Laboratory Hub (2009-2021), with clinical testing (single gene/whole exome) of 158 suspected HLRCC and 952 Renal Cell Carcinoma (RCC) patients. The Sheffield/Leeds datasets comprised per-patient data from probands with RCC or suspected HLRCC referred to the North East and Yorkshire Genomic Laboratory Hub (2016-2021), with clinical testing (single gene/multi-gene panel) of 229 suspected HLRCC and 131 RCC patients (Sheffield) and 359 RCC patients (Leeds). The Ambry dataset comprised per-patient data from clinical testing for *FH* undertaken at Ambry Genetics of 338 patients referred for kidney cancer genetic testing from US clinical genetics and endocrinology centres (2013-2017). In total, data from 387 suspected HLRCC and 1780 renal cancer probands were available for analysis.

At a minimum, data collected contained (i) GRCh37 *FH* variant HGVS nomenclature, (ii) patient phenotype, gene panel used, and/or referral reason (see Supplementary Methods)(7). Where available, variant data from individuals with a renal cancer subtype was noted. Individuals in whom *FH* testing was for the biallelic Fumarase Deficiency (FMRD) phenotype were not included(8).

### Assembly of control data series

Population controls were collated from three population datasets: the 1000 Genomes Project (1KGP), comprising data from 2504 individuals across 5 top-level ancestries (African (n=661), Admixed American (n=347), East Asian (n=504), European (n=503), South Asian (n=489)); gnomAD v2.1.1 non-cancer exome data, comprising data from 118,474 individuals (African/African American (n=7,451), Admixed American (n=17,130), Ashkenazi Jewish (n=4,786), East Asian (n=8,846), Finnish Europeans (n=10,816), Non-Finnish Europeans (n=51,372), South Asian (n=15,263), Remaining individuals (n=2,810)); and UK Biobank, comprising data from 441,317 individuals across 6 top-level ethnicities (White (n=417,249), Asian (n=8,872), Black (n=7,143), Chinese (n=1,387), Mixed (n=2,614), Other (n=4,052))(9–11). UK Biobank participants were additionally filtered to include only non-cancer participants (Supplementary Methods). Acknowledging that there is some minor overlap between the 1KGP phase 3 data and gnomAD v2.1.1, both datasets were included to increase representation of non-white ancestries.

### Defining missense variants and truncating variants

Using annotations provided by Ensembl VEP against the MANE Select transcript for *FH* (NM_000143.3), *FH* variants in cases and controls were further categorised into truncating variants (annotations containing frameshift_variant, stop_gained, splice_acceptor_variant, and/or splice_donor_variant) and missense variants (missense_variant and missense_variant&splice_region_variant) (12).

The final variant sets were run through VariantValidator against the same *FH* transcript(13). For each missense variant identified, *in silico* prediction scores were generated using REVEL and BayesDel scores modelled without allele frequency (noAF)(14, 15). ClinVar assertions and LOVD classifications were obtained for each missense variant, with each database searched on 07/10/2024(16, 17).

### Calculation of predicted maximum tolerated allele frequency (MTAF)

To calculate a population frequency threshold below which variants may be described as “very rare”, a Maximum Tolerated Allele Frequency (MTAF_pred_) was calculated using the methods described by Whiffin et al(18, 19). Namely, for a monoallelic phenotype, MTAF_pred_ = 0.5 x disease prevalence x (genetic heterogeneity x allelic heterogeneity) x 1/(penetrance).

**Disease prevalence:** National incidence rates of renal cancer vary widely across the world(20). As such, given the majority of the datasets used in this paper originated from UK testing laboratories, we used an estimate of lifetime risk of the disease of 1 in 50 based on average estimates in the UK population of frequency in males (1 in 38) and females (1 in 68)(21).

**Disease penetrance:** The estimated penetrance of renal cell cancer for *FH* variant carriers varies widely between publications and depending on ascertainment, with one publication based on data from ExAC reporting penetrance to be 1.7-5.8%, and others based on HLRCC cohorts stating the risk of developing renal cell cancer in *FH* variant carriers to be 15-34%, varying in reporting penetrance to age 40 versus lifetime penetrance(1, 22, 23). An intermediate estimate of penetrance of 15% for lifetime risk of renal cell carcinoma was used for the MTAF_pred_ calculation.

**Genetic and allelic heterogeneity:** A genetic heterogeneity estimate of 0.0022 (0.22%) was used based on 3/1336 identified P/LP *FH* variants in a cohort of RCC patients(24). As previously described for the *SDHB* and *SDHD* genes, a conservative estimate of 0.1 was applied for allelic heterogeneity (the proportional contribution of FH variant of uncertain penetrance under evaluation)(18).

From these parameter estimates, an MTAF_pred_ of 1.5×10^-5^ was estimated. We computed a maximum tolerated allele count (MTAC) using the MTAF_pred_ for each ethnicity group in each population control dataset as described by Whiffin *et al*., derived from the upper 95% confidence interval of the underlying Poisson distribution and assuming adequate coverage(19). A very rare missense variant (VRMV) or very rare truncating variant (VRTV) was thus defined as a variant for which the allele count in controls did not exceed the MTAC in any individual ethnicity group. Analyses were repeated using the MTAC lower 95% confidence interval to explore the impact of a stricter definition of rarity (Supplementary Methods, Supplementary Table 1).

Calculation of pan-gene very rare variant likelihood ratios (PG-VRV-LRs) As per the methods described in Garrett et al. 2022, we generated positive LRs and confidence intervals based on the rate of the variant under study in ‘positive’ cases of the relevant phenotype (true positive rate) compared with the rate of variant entity under study in those negative for the relevant phenotype i.e. population controls (false positive rate). The positive LR = (a/a + c)/(b/b + d), where a = true positive, b = false positive, c = false negative, and d = true negative(18).

The PG-VRV-LRs were generated for each of VRMVs and VRTVs found in each of HLRCC and renal cancer cases compared with general population controls (1KGP, gnomAD v2.1.1 (non-cancer), and UK Biobank). These positive LRs were converted into log likelihood ratios (LLRs) equivalent to the widely adopted ACMG/AMP “evidence points” using a log base 2.08 conversion as prescribed in the Bayesian update of the ACMG/AMP variant classification framework described by Tavtigian et al., 2020(25). Calculations were repeated with (i) removal from the renal cancer testing cohort where pathology is known to be papillary-type renal cell cancers (as individuals with this renal subtype may be eligible for inclusion as HLRCC) and (ii) removal of variants ascribed as pathogenic or likely pathogenic in ClinVar (to appraise the evidence estimate applicable to new variants of uncertain pathogenicity, thus excluding these (typically recurrent) variants of established pathogenicity).

### Calculation of domain-specific very rare missense variant likelihood ratios (DS-VRMV-LRs)

Using the clustering algorithm and windowing method as described by Walsh et al.(26), we examined regional enrichment of VRMVs in cases compared to VRMVs in controls (DS-VRMV-LRs).

## Results

### Identification of Very Rare Variants in *FH* in cases and population controls

In total, the data from two laboratories in whom testing of *FH* was performed on the basis of an NHS testing eligibility of HLRCC (‘HLRCC cases’) comprised 387 individuals. Of these, a very rare truncating variant (VRTV) in *FH* was identified in 79/387 (20.41%) HLRCC cases, with 42 unique VRTVs identified. A very rare missense variant (VRMV) in *FH* was identified in 146/387 (37.73%) non-overlapping HLRCC cases, with 61 unique VRMVs identified. For these analyses, a ‘very rare’ variant was defined as being of a sufficiently low frequency to be consistent with pathogenicity within all control populations analysed (see Methods).

In total, the data from four laboratories in whom testing of *FH* was performed in individuals with renal cancer as part of a broader renal cancer panel comprised 1,780 renal cancer cases. Of these, an *FH* VRTV was identified in 13/1,780 (0.73%) and a VRMV in *FH* was identified in 18/1,780 (1.01%).

The three population datasets comprised 562,295 individuals in total: 469,124 individuals were described as being of white/non-Finnish European ancestry and 93,171 individuals were of other ancestries (see Methods). The frequency in the amalgamated population controls was 43/562,295 (0.0076%) for *FH* VRTVs and 988/562,295 (0.18%) for *FH* VRMVs across all ancestries; restricting to white population controls, the frequency was 36/469,124 (0.0077%) for *FH* VRTVs and 802/469,124 (0.17%) for *FH* VRMVs.

**Table 1:** Summary of tested individuals and rare *FH* variants identified across the four laboratories, and observations in controls from gnomAD v2.1.1, the 1000 Genomes Project, and UK Biobank. 1KGP, 1000 Genomes Project; EUR, European; HLRCC, Hereditary.

|  | HLRCC |  |  | Renal Cancer |  |  |  |  | White Ethnicity Population Controls |  |  |  | All Population Controls |  |  |  |
| --- | --- | --- | --- | --- | --- | --- | --- | --- | --- | --- | --- | --- | --- | --- | --- | --- |
|  | Sheffield | Birmingham | Total | Sheffield | Leeds | Birmingham | Ambry | Total | gnomAD NFE | 1KGP EUR | UKB White | Total | gnomAD all | 1KGP all | UKB all | Total |
| Rare truncating | 46 | 33 | 79<br>(20.4%) | 1 | 6 | 5 | 1 | 13<br>(0.73%) | 6 | 0 | 30 | 36<br>(0.0077%) | 12 | 0 | 31 | 43<br>(0.0076%) |
| Rare missense | 86 | 60 | 146<br>(37.7%) | 0 | 2 | 10 | 6 | 18<br>(1.01%) | 117 | 0 | 685 | 802<br>(0.17%) | 240 | 0 | 748 | 988<br>(0.18%) |
| Total individuals tested | 229 | 158 | 387 | 131 | 359 | 952 | 338 | 1780 | 51372 | 503 | 417249 | 469124 | 118474 | 2504 | 441317 | 562295 |

**Leiomyomatosis and Renal Cell Cancer; NFE, Non-Finnish European; UKB, UK Biobank.**

### Calculation of Likelihood Ratios and Log Likelihood Ratios

For the HLRCC phenotype, the pan-gene very rare variant likelihood ratio (PG-VRV-LR) for *FH* VRTVs was 2,669.39 (79/387 in HLRCC cases versus 43/562,295 in population controls), which equates to a log likelihood ratio base 2.08 (LLR) of 10.77. The PG-VRV-LR for *FH* VRMVs was 214.71 (146/387 in HLRCC cases versus 988/562,295 in population controls), which equates to a LLR of 7.33.

In the group of renal cancer cases, the corresponding figures were a likelihood ratio for *FH* VRTVs of 95.50 (13/1,780 in renal cancer cases versus 43/562,295 in population controls), equating to a LLR of 6.23. The PG-VRV-LR was 5.76 for *FH* VRMVs, (18/1,780 in renal cancer cases versus 988/562,295 in population controls), which equates to a LLR of 2.39 (Table 2, Supplementary Table 2).

**Table 2:** Likelihood Ratio calculations for PG-VRV-LRs in the HLRCC and unselected renal cancer phenotypic groups. ACMG evidence points (EPs) calculated by log conversion to base 2.08 of each likelihood ratio(25). HLRCC, Hereditary Leiomyomatosis and Renal Cell Cancer; LLR, log-likelihood ratio; LR, likelihood ratio; P/LP, Pathogenic/Likely Pathogenic.

|  |  | Cases with variant | Controls with variant (/562,295 controls) | LR (95% Confidence Intervals) | LLR |
| --- | --- | --- | --- | --- | --- |
| **HLRCC**<br>(/387 cases) | Very Rare Truncating Variants | 79 | 43 | 2669.39<br>(1843.44-3881.25) | 10.77 |
|  | Very Rare Missense Variants (VRMV) | 146 | 988 | 214.71<br>(185.01-246.87) | 7.33 |
|  | Very Rare Missense Variants (VRMV), 2-star P/LP removed | 46 | 914 | 73.13<br>(54.44-97.01) | 5.86 |
|  | Very Rare Missense Variants (VRMV) 1-star and 2-star P/LP and conflicting removed | 30 | 754 | 57.81<br>(39.79-82.71) | 5.54 |
| **Renal**<br>(/1780 cases) | Very Rare Truncating Variants | 13 | 43 | 95.50<br>(48.93-183.0) | 6.23 |
|  | Very Rare Missense Variants (VRMV) | 18 | 988 | 5.76<br>(3.51-9.30) | 2.39 |
|  | Very Rare Missense Variants (VRMV), 2-star P/LP removed | 7 | 914 | 2.42<br>(1.06-5.23) | 1.21 |
|  | Very Rare Missense Variants (VRMV), 1-star and 2-star P/LP and conflicting removed | 4 | 754 | 1.68<br>(0.54-4.61) | 0.71 |

The PG-VRV-LR was also calculated for each phenotype using a stricter definition for very rare variant inclusion (Supplementary Methods, Supplementary Table 3). The PG-VRV-LR for HLRCC *FH* VRTVs increased from 2,669.39 to 4,615.28, or a slightly increased LLR of 11.52. Similarly, the PG-VRV-LR for HLRCC *FH* VRMVs increased from 214.71 to 642.66, which equates to an LLR of 8.83. The same impact was seen for renal cancer cases, where the PG-VRV-LR for *FH* VRTVs increased from 95.50 to 185.82, equating to an LLR of 7.13, and the PG-VRV-LR for FH VRMVs increased from 5.76 to 18.23, equating to an LLR of 3.96.

### Sensitivity analyses for Likelihood Ratios

#### Modification of control population

To further test for sensitivity, calculation of likelihood ratios was repeated with controls restricted to individuals of white ethnicity, as the narrower ethnicity status would constitute a more standardised reference group for comparison (Supplementary Table 2). The frequencies of variants altered very modestly: from 0.176% in all controls to 0.171% in white ethnicity controls for VRMVs and from 0.00765% to 0.00767% for VRTVs. For the HLRCC phenotype, the pan-gene likelihood ratio for *FH* VRTVs was adjusted minorly to 2736.12, equating to an LLR of 10.81, and the likelihood ratio for *FH* VRMVs was 220.68, equating to an LLR of 7.37. For renal cancer cases, the pan-gene likelihood ratio for *FH* VRTVs was 97.89, equating to an LLR of 6.26, and for *FH* VRMVs the likelihood ratio was 5.92, equating to an LLR of 2.43 (Supplementary Table 2).

#### Consolidation of inclusion of renal cancer subtypes

A person with papillary renal cancer may be included in both the HLRCC and renal cancer cohorts. There is therefore a possibility of this highly-specific histology inflating the renal cancer PG-VRV-LRs, making these LRs not as relevant in the context of other renal cancer sub-types. To investigate this possibility, the analysis for the renal cancer group was repeated with removal of individuals in whom the histology was actively documented as being papillary renal cell cancer (Supplementary Methods; Supplementary Tables 2 and 4). For VRTVs, this resulted in modest reduction of the pan-gene likelihood ratio to 92.50 (11/1,555 renal cancer cases versus 43/562,295 population controls), equivalent to an LLR of 6.18, and for VRMVs the likelihood ratio was 5.86 (16/1,555 renal cancer cases versus 988/562,295 population controls), equating to an LLR of 2.41 (Supplementary Table 2).

#### Exclusion of previously-reported pathogenic variants

Several *FH* missense variants in the case series had been previously reported as Likely Pathogenic or Pathogenic within ClinVar. To avoid possible enrichment on account of frequently-observed well-established pathogenic variants from the data (and limit the possibility that association between phenotype and VRMVs was driven by one or two common pathogenic variants), likelihood calculations were again repeated with removal of variants reported in ClinVar as Likely Pathogenic or Pathogenic. We first removed from both case and control data variants assigned 2-star review status (i.e. reported and consistently classified by at least two laboratories). We then removed variants assigned as 1-star or 2-star, or with a ‘conflicting’ assertion involving one or more (likely) pathogenic classifications. As might be anticipated, exclusion of these Pathogenic/Likely Pathogenic variants resulted in sizeable diminution of the LRs (Table 2, Supplementary Tables 2, 5, and 6).

### Alignment with *in silico* prediction tools

We annotated each missense variant with two commonly used *in silico* prediction tools (Supplementary Tables 5-6). Using predictions from REVEL and BayesDel (no AF), we applied the thresholds described by the ClinGen Sequence Variant Interpretation (SVI) group in Pejaver et al., 2022 to determine the evidence strengths that could be afforded for each variant(14, 15, 27). Using these thresholds, 61/61 VRMVs in HLRCC and 18/18 VRMVs in renal cancer would qualify for at least supporting evidence using either REVEL or BayesDel, with 60/61 and 16/18 attaining at least moderate evidence.

### Calculation of domain-specific Likelihood Ratios in HLRCC

Following spatial clustering analysis in genomic space of VRMVs comparing HLRCC cases against controls, three ‘hotspot’ domains were identified as being enriched for VRMVs in HLRCC cases compared to population controls (p.107-117, P-value 1.69×10^-16^; p.230-234, P-value 3.67×10^-26^; p.365-397, P-value 2.16×10^-7^). These three domains together comprised 9% of the total protein length (Figure 1A). 81/146 VRMVs found in HLRCC cases lay within the clustered regions compared to 96/2264 in controls. Thus, in comparison to the pan-gene likelihood ratio for *FH* VRMVs of 214.71 (LLR 7.33), the domain-specific likelihood ratio for *FH* VRMVs (DS-VRMV-LR) for these three regions taken together was increased to 1225.93 (LLR 9.71), whilst outside of these domains the DS-VRMV-LR was commensurately reduced to 105.88, equivalent to an LLR of 6.37 (Supplementary Table 7). Examination of the variants at these hotspots revealed that there was significant contribution from the established, recurrent (likely) pathogenic variants (Figure 1A). A second run of the spatial clustering analysis following removal of established (likely) pathogenic variants (defined as 1-star or 2-star P/LP variants in ClinVar, including conflicting classifications) revealed an adjusted single region comprising 28.5% of the protein (p.277-408, P-value 8.72×10^-5^; Figure 1B). The DS-VRMV-LR when recurrent variants were removed was 166.37 for VRMVs falling within this adjusted domain (an LLR of 6.98), and 34.98 for VRMVs outside this domain (an LLR of 4.85), with the corresponding pan-gene metrics being an LR of 57.81 and LLR of 5.54.

**Figure 1:**
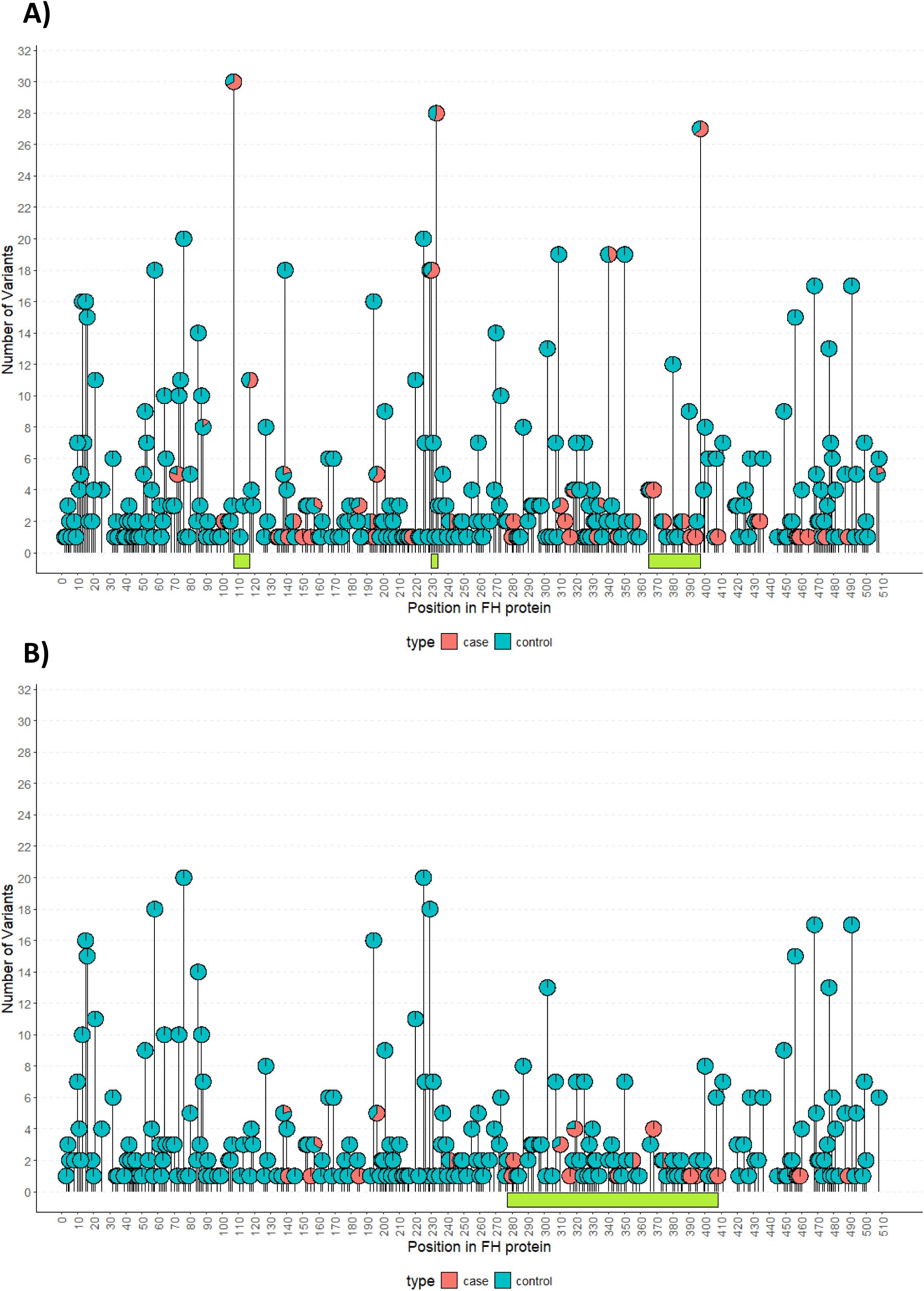
Lollipop plot showing the position of *FH* VRMVs in A) all 387 HLRCC cases and 562,295 population controls, and B) HLRCC cases and population controls after individuals with recurrent P/LP VRMVs were removed. Variants identified in HLRCC cases are highlighted in red and variants identified in population controls are highlighted in blue. Boundaries for the variant hotspot region(s) identified using a clustering algorithm (see Methods) are shown beneath each plot in green.

## Discussion

We calculated a maximum tolerated allele frequency (MTAF) threshold that we used to ascribe variants as being “very rare”, that is of sufficient rarity in the population to be plausibly pathogenic. Using data from sequencing of *FH* in 387 cases of HLRCC and 562,295 population controls, we have demonstrated that identification of an *FH* VRMV is 214.7-fold more likely in the HLRCC case group than in the general population. Even removing the VRMVs that are already previously reported as (Likely) pathogenic in ClinVar (1-star, 2-star and conflicting classifications), this likelihood ratio remains sizeable at 57.81. For comparison, undertaking this analysis for protein truncating variants, the likelihood ratio was 2,669.39.

By comparison and as might be anticipated, the likelihood ratios in the context of renal cancer are more modest due to the lower specificity for this phenotype. The likelihood ratio for *FH* VRTVs for all renal cancer was 95.50. The likelihood ratio for *FH* VRMVs for all renal cancers was 5.76, falling to 1.92 following removal of the established, previously-reported ClinVar (likely) pathogenic variants (and cases of known papillary renal cell cancer).

Appropriate weighting of phenotypic data for clinical variant classification has long been problematic.

The default allocation in the 2015 ACMG/AMP framework of ‘supporting’ evidence for PP4 was deliberately cautious. This was in response to previous ascertainment bias in ascribing of rare variants as being (likely) pathogenic purely or primarily on the basis of observing the variant in an individual with disease. For instances where the phenotype was comparatively common and only modestly attributable to a given gene, this would typically represent gross over-weighting of phenotypic specificity. For example, observation of a *BRCA1* missense variant in a case of breast cancer, even if of younger onset. This over-weighting for phenotypic specificity had been especially problematic in the era predating availability of large control datasets by which we can now better assess variant frequencies in the population and (lack of) constraint for missense variants. However, the cautious scoring for PP4 allocated within the ACMG/AMP framework has resulted in uncertainty and possibly excess conservatism around allocation of evidence in the opposite scenario, namely where the phenotype is very rare, the phenotype is highly specific for that gene and large population datasets can confirm the variant as extremely rare.

We previously examined the rare endocrine tumour entity of phaeochromoytoma/paraganglioma (PHAEO/PGL) and VRMVs in *SDHB* (and subsequently *SDHD*) in order to explore quantitation of phenotypic specificity for a given gene/phenotype/variant-class relationship(18). Following definition of the corresponding MTAFs, using laboratory-testing data for 6,328 cases of PHAEO/PGL, we demonstrated a VRMV-LR for *SDHB* compared to population controls of 76.2 (54.8-105.9), which after removal of the recurrently reported pathogenic VRMVs was reduced to 34.5 (24.2-49.1), which corresponded to log_2.08_LRs of 5.92 and 4.83 respectively. We were able to make use of a stand-alone, smaller, well-characterised research series of PHAEO/PGL, in which via case-only logistic regression we demonstrated that *SDHB* and *SDHD* PVs were more associated with head and neck paragangliomas (HNPGL, compared to sympathetic PHAEO/PGL), with invasive disease (compared to non-invasive disease), with multiple tumours (versus solitary tumours) and with familial disease (versus isolated disease).

Nevertheless, we lacked information regarding these detailed clinical attributes for the large laboratory-testing series in which we had calculated the empirical VRMV-LRs. Thus, whilst presence/absence of a PHAEO/PGL would appear in principle a straight-forward binary phenotype, there is heterogeneity in regard of the “strength of phenotype” as relates to these characteristics of underlying genetic enrichment.

The HLRCC cases will of course also be heterogeneous in regard of the “strength of phenotype”. Our HLRCC case definition required the tested proband to have Type 2 papillary OR tubulo-papillary renal cancer, cutaneous leiomyoma and/or uterine leiomyoma with stipulations regarding severity of features, age-of-onset, histology and family history (Box 1), as per NHS testing eligibility. Although clinicians ordering diagnostic genetic testing within an NHS clinical genetics centre can reasonably be assumed to adhere to eligibility requirements, the diagnostic testing laboratories were unable to provide detailed descriptions for each proband of the phenotypic elements present. The consequence, if the phenotype of some of the included cases fell below eligibility criteria, would be that the LRs presented will have been underestimated. However, conversely, the eligibility criteria represent the lower threshold for inclusion: some included cases may have had personal and family histories that exceeded this bar. Thus we do not know the “average” level of HLRCC phenotype, only the prescribed minimum. Accurate phenotyping for pleiomorphic disorders is highly challenging even when undertaken for well characterised research series. Even where there is detailed documentation of the phenotypic features present, comprehensive imaging has rarely been performed by which to actively exclude presence of other features (emergence of which may also be age-dependent)

Thus, whilst we are able to present a net likelihood ratio (measure of enrichment) applicable to the group meeting this definition of HLRCC, we were unable to provide more granular scoring on the basis of individual phenotypic elements or combinations. To use these data to inform allocation of PP4 for clinical variant interpretation, the VRMV likelihood ratio might serve as a baseline evidence weighting for a standard or average HLRCC case that meets these eligibility criteria. Up- or down-weighting may then be advised in the context of phenotypes at the milder or more severe end of the qualifying eligibility spectrum (assuming phenotypic severity would take a Galton-type (left-skewed) distribution).

We also use the distribution of VRMVs across the gene to define an enriched cluster region, where the likelihood ratio for VRMVs is further enhanced in comparison to the ‘colder’ region of the gene: again, this information is potentially useful to inform differential allocation of phenotype-based evidence points, depending on the region of the gene in which a new missense variant is encountered (thus effectively bringing in PM1-domain weighting).

## Limitations

The primary limitation to this analysis, as already described and common for clinically ascertained series, was the lack of detailed individual-level phenotype data, meaning we were unable to partition the HLRCC group by clinical features/severity. Additionally, in regard of incomplete individual level phenotype data for the renal cancer series, we could not evaluate the extent to which the range of pathologies was representative. Furthermore the quoted likelihood ratio may still be inflated due to contamination with papillary renal cell carcinomas.

There were some further limitations to this analysis. Firstly, we lacked data on ancestry for the case series, which represented the HLRCC and renal cancer patient populations tested across three UK and one US genetics laboratories. The cases would likely be of predominantly Western European ancestry but with a sizeable minority of other ancestries. The primary issue here is that our assignation of variants as being very rare is of lesser confidence for non-Western European ancestry groups. Firstly, the smaller size of the non-white populations within control datasets will inherently limit the precision by which we can estimate variant frequencies and assign the cut-off as very rare. Variants with a true frequency above the MTAF_pred_ in that ancestry group may, by chance, still be absent in these modest-sized control series (albeit that we did have three different data sources with multiple non-Western European ancestry groups). Secondly, although unlikely due to the diverse groups included in particular within the 1KGP, it is possible that there may be HLRCC cases from ancestry groups wholly not represented in any control series, meaning again we may erroneously have included some variants as being ‘very rare’ that are comparatively common in that case’s ancestry group. Erroneous inclusion of such variants as VRMVs in the case series may result in overestimation of the PG-VRMV-LRs.

An additional limitation, as applicable to most rare syndrome gene scenarios, is the accuracy of parameter estimates for the MTAF estimation: published estimates for lifetime penetrance and genetic heterogeneity vary widely and may be subject to substantial ascertainment bias. A further limitation (or consideration) is that the VRMV-LR metrics are based on and applicable only to observed variants of presumptive full penetrance that themselves are very rare (i.e. are observed in the control population at frequencies below the MTAF_pred_). Variants that are disease-associated but of lower penetrance will likely occur in the population but at frequencies above the MTAF_pred_. Such reduced penetrance variants would not have been included in the VRMV case control analyses, and the VRMV-LR metric would not be applicable to them.

## Conclusions

In summary, we present data comparing against control populations the frequency of very rare *FH* missense variants detected on clinical diagnostic testing in (i) the highly specific phenotypic context of HLRCC, contrasting this to (ii) the much less specific phenotypic context of renal cancer. We have quantified the enrichment as a likelihood ratio, which can be converted into a log likelihood ratio amenable to incorporation in the Bayesian update of the ACMG/AMP variant classification framework. We have also demonstrated how this enrichment varies for different regions of the gene, presenting ‘domain-specific’ likelihood ratios. These data are of potential clinical utility for quantifying evidence allocation for PP4 (and PM1) when an *FH* variant is observed in an individual with this rare and highly specific phenotype.

## Supporting information

Supplementary Note: CanVIG-UK Consortium

## Data Availability

All data produced in the present work are contained in the manuscript.

## Acknowledgements

A full list of CanVIG-UK consortium members and their affiliations appears in the Supplemental Material.

## Funding statement

S.A. and C.F.R. are supported by CG-MAVE, CRUK Programme Award [EDDPGM-Nov22/100004]. A.G. and H.H. are supported by CRUK Catalyst Award CanGene-CanVar (C61296/A27223). E.R.W. is supported by the Manchester NIHR Biomedical Research Centre (IS-BRC-1215-20007).

## Author contributions

Conceptualization: C.T.; Data Curation: S.B., M.D., C.H., R.R.; Formal Analysis: S.A.; Funding Acquisition: C.T.; Methodology: C.T., E.R.M., A.A., T.M.V., A.G., M.D., G.J.B., A.C., J.F., B.F., S.P.S., J.G., J.P., T.M.D., K.S., H.H.; Project administration: S.A.; Visualization: S.A..; Writing-original draft: C.T., S.A.; Writing-review and editing: All authors

## Ethics Declaration

The human variant data used were all de-identified. The data used for frequency analyses were provided in summary form only; all these data were wholly de-identified, and thus, institutional review board approval was not required.

## Conflicts of interest

The authors declare that they have no competing interests.

## Availability of data and materials

All data generated or analysed during this study are either included in this published article and its supplementary information files, are available from the corresponding author on request, or are publicly available at the references and URLs provided. This research has been conducted using the UK Biobank Resource under Application Number 76689 (https://www.ukbiobank.ac.uk/).

## Notes

### Competing Interest Statement

The authors have declared no competing interest.

### Author Declarations

This work did not require IRB or ethics committee approvals. The human variant data used were all de-identified. The data used for frequency analyses were provided in summary form only; all these data were wholly de-identified, and thus, institutional review board approval was not required.

