## Supplementary Methods for "Quantifying evidence for phenotypic specificity (PP4) for syndromic phenotypes: Large-scale integration of rare germline *FH* variants from diagnostic laboratory testing for HLRCC (Hereditary Leiomyomatosis and Renal Cell Cancer) and renal cancer"

### **Supplementary Materials**

#### **Supplementary Methods**

##### **Data Cleaning and Filtering**

Per-patient summary level data from unrelated patients referred for *FH* testing was collated from three UK diagnostic clinical genetics laboratories (Birmingham, Leeds, Sheffield) and one US-based central testing laboratory (Ambry):

- Birmingham: 1110 Patients referred to the West Midlands Regional Genomic Laboratory Hub between 2009-2021
- Leeds: 359 Patients referred to the Leeds Genetics Laboratory within the North East and Yorkshire Genomic Laboratory Hub between 2016-2021
- Sheffield: 360 Patients referred to the Sheffield Diagnostic Genetics Service within the North East and Yorkshire Genomic Laboratory Hub between 2016-2021
- Ambry: 338 Patients referred to Ambry Genetics between 2013-2017

Based on the information available from each laboratory, HLRCC and Renal cancer phenotypes were defined as such:

- HLRCC: Diagnosis, pathology, or referral details state HLRCC, and/or renal papillary cancer (type II or unknown) including bilateral renal papillary cancer. Records reporting cutaneous leiomyomata and/or uterine leiomyomata as the reason for testing must also include HLRCC or kidney cancer pathology, as family history and histology was not available for most variant data, and leiomyomata alone is not sufficient for eligibility for R365 testing.
- Renal: Diagnosis, pathology, or referral details state kidney or renal cancer of any sub-type as the reason for testing. If none of these details were available, individuals were included if they were tested using the R224 renal cancer gene panel or laboratory-specific renal cancer diagnostic gene panel.

Patients with a reported Fumarate Hydratase Deficiency (FHD) phenotype were not included in either dataset due to risk of penetrance of these *FH* variants only being significant for FHD and no other phenotype. Individuals with an unknown test indication or a test indication not matching either phenotype were not included.

For all cases, the test indication indicated diagnostic testing rather than familial/predictive testing. Where variant data was provided as genomic coordinates (GRCh37) or the corresponding LRG coordinates (LRG_504), variants were converted to HGVS nomenclature using VariantValidator^1^ against the MANE Select transcript for *FH* (RefSeq: NM_000143.3).

##### **Exclusion of renal papillary cancer cases**

When removing individuals with known renal papillary cancer from the dataset, records were excluded if the diagnosis, pathology, or referral details stated renal papillary cancer of any sub-type. In the Ambry dataset, no records contained referral details with any HLRCC phenotype except renal papillary cancer, therefore the Ambry HLRCC dataset is roughly equivalent to a renal papillary phenotype dataset. In the Birmingham dataset, there were no reported renal papillary cancer cases. This does not mean there were no renal papillary cancer cases, only that there were none reported in the dataset containing variant information.

##### **Filtering of population controls**

Population controls were derived from the 1000 Genomes Project (1KGP) phase 3 data (total individuals: 2504), the gnomAD v2.1.1 non-cancer exomes dataset (total individuals: 118474), and a static download from the UK Biobank from September 2022 (total individuals: 441317)^2-4^. Data from the 1KGP was included to increase representation from rarer ethnicities. Data from gnomAD v2.1.1 non-cancer exomes was filtered to remove variant observations which failed the standard gnomAD quality control filters. UK Biobank data was processed and filtered as follows:

- Downloaded initial BED file listing the coordinates of all exonic regions +/- 25 bp plus UTRs in the build 38 GENCODE v41 annotation across all FH transcripts.
- A cohort of 374,363 “non-cancer” patients was defined by applying the following filters to the bulk participant data encompassing just over 500,000 individuals:
  - “Cancer diagnosed by doctor” = “No”
  - “Cancer code, self reported” is NULL
  - “Type of cancer: ICD9” is NULL
  - “Type of cancer: ICD10” is NULL
- Of the non-cancer patients, 330,652 had had exome sequencing conducted, and all variants overlapping any of the FH coding regions were extracted for these samples.
- FH variants were lifted back to GRCh37 using UCSC liftOver^5^
- For each variant in the resulting VCF, the total allele number (AN) and total variant allele count (AC; adding 1 for heterozygous carriers and 2 for homozygous carriers) was generated by looping through each participant and each ethnicity.
- Participants were included only if they listed a single ethnicity to enable later grouping by ethnicity. Ethnicities of ‘Prefer not to answer’ and ‘Do not know’ were excluded from analysis. The ethnicity groupings and sub-groupings can be seen at <https://biobank.ctsu.ox.ac.uk/crystal/field.cgi?id=21000>.
- All variants were finally run through VEP web browser, Ensembl 108 to retrieve the HGVSc/p coordinates and VEP annotations^6^.

##### **MTAF_pred_ and defining very rare variants**

The Maximum Tolerated Allele Frequency (MTAF_pred_) was calculated as defined previously^7,8^:

$${MTAF}_{pred} = v \left( ga \right) \frac{1}{2p}$$

where:

- $v$ = disease prevalence
- $g$ = genetic heterogeneity
- $a$ = allelic heterogeneity
- $p$ = penetrance

Using the Whiffin/Ware allele calculator, we computed the maximum tolerated allele count (MTAC) given the MTAF_pred_ for *FH* for each top-level ethnicity group in the population control datasets and assuming a Poisson distribution and estimates based on the upper 95% confidence interval (Supplementary Table 1). A variant which is present at an allele frequency below the MTAC for each ethnicity in each population dataset was considered to be very rare, a truncating variant which was very rare denoted as a Very Rare Truncating Variant (VRTV), and a missense variant which was very rare denoted as a Very Rare Missense Variant (VRMV).

The top-level ethnicity backgrounds were used to group multiple sub-ethnicities from UK Biobank as per data field specification 21000; this is to avoid where possible a 0-vs-1 inclusion for maximum tolerated alleles in the sub-populations (where a single observation in a small subpopulation would mean the variant is not considered as a very rare missense variant). The Chinese AN is the only top-level group where 1 observation would affect designation of VRMV, however all variants with observations in this sub-population exceed the Maximum Tolerated Allele Count (MTAC) in at least one other sub-population or in the total dataset.

The MTAC was also re-computed for each ethnicity group in the population control datasets using the lower 95% confidence interval of the underlying Poisson distribution (Supplementary Table 1). This was to generate very rare variant groups based on a stricter definition of ‘very rare’ and examine the impact of this on the resulting likelihood ratio calculations.
