## Supplementary Note: CanVIG-UK Consortium for "Quantifying evidence for phenotypic specificity (PP4) for syndromic phenotypes: Large-scale integration of rare germline *FH* variants from diagnostic laboratory testing for HLRCC (Hereditary Leiomyomatosis and Renal Cell Cancer) and renal cancer"

**Supplementary Note 1: CanVIG-UK Consortium Members and Affiliations**

C. Turnbull^1,49^, A. Garrett^1,47^, L. Loong^1^, S. Choi^1^, B. Torr^1^, S. Allen^1^, M. Durkie^2^, A. Callaway^3^, J. Drummond^4^, G.J. Burghel^5^, R. Robinson^6^, I.R. Berry^65^, A.J. Wallace^5^, D.M. Eccles^7, 8^, M. Tischkowitz^13^, S. Ellard^9^, H. Hanson^1,16^, E. Baple^10,11^, D.G. Evans^5,30^, E. Woodward^5,30^, F. Lalloo^5,30^, S. Samant^33^, A. Lucassen^57,14,15^, A. Znaczko^44^, A. Shaw^23^, A. Ansari^34^, A. Kumar^21^, A. Donaldson^53^, A. Murray^19^, A. Ross^18^, A. Taylor-Beadling^22^, A. Taylor^18^, A. Innes^25^, A. Brady^29^, A. Kulkarni^23^, A.C. Hogg^5,^ A. Ramsay Bowden^18^, A. Hadonou^47^, B. Coad^16^, B. McIldowie^19^, B. Speight^18^, B. DeSouza^47^, B. Mullaney^3^, C. McKenna^62^, C. Brewer^44^, C. Olimpio^18^, C. Clabby^40^, C. Crosby^47^, C. Jenkins^42^, C. Armstrong^33^, C. Bowles^9^, C. Brooks^22^, C. Byrne^62^, C. Maurer^4^, D. Baralle^57^, D. Chubb^1^, D. Stobo^34^, D. Moore^35^, D.O'Sullivan^33^, D. Donnelly^62^, D. Randhawa^24^, D. Halliday^41^ , E. Atkinson^50^, E. Rauter^24^, E. Johnston^38^, E. Maher^8^, E. Sofianopoulou^17^, E. Petrides^23^, F. McRonald^43^, F. Pelz^51^, I. Frayling^19^, G. Corbett^62^, G. Rea^62^, H. Clouston^5^, H. Powell^31^, H. Williamson^52^, H. Carley^47^, H.J.W. Thomas^26^, I. Tomlinson^63^, J. Cook^46^, J. Tellez^32^, J. Whitworth^18^, J. Williams^49^, J. Murray^35^, J. Campbell^27^, J. Tolmie^33^, J. Field^38^, J. Mason^64^, J. Burn^31^, J. Bruty^18^, J. Callaway^8^, J. Grant^34^, J. Del Rey Jimenez^47^, J. Pagan^35^, J. VanCampen^24^, J. Barwell^53^, K. Monahan^29^, K. Tatton-Brown^16,^ K.R. Ong^63^, K. Murphy^33^, K. Andrews^18,^ K. Mokretar^23^, K. Cadoo^48^, K. Smith^52^, K. Baker^8^, K. Brown^24^, K. Reay^64^, K. McKay Bounford^34^, K. Bradshaw^38^, K. Russell^65^, K. Stone^23^, K. Snape^16^, L. Crookes^5^, L. Reed^21^, L. Yarram^65^, L. Cobbold^47^, L. Walker^39^, L. Walker^41^, L. Hawkes^16^, L. Busby^22^, L. Izatt^23^, L. Kiely^22,^ L. Hughes^64^, L. Side^56^, L. Sarkies^18^, K.-L. Greenhalgh^28^, M. Shanmugasundaram^63^, M. Duff^40^, M. Bartlett^29^, M. Watson^3^, M. Owens^9^, M. Bradford^54^, M. Huxley^64^, M. Slean^33^, M. Ryten^23^, M. Smith^55^, M. Ahmed^21^, N. Roberts^2^, O. Middleton^33^, P. Tarpey^4^, P. Logan^62^, P. Dean^3^, P. May^24^, P. Brace^21^, R. Tredwell^38^, R. Harrison^37^, R. Hart^63^, R. Kirk^5^, R. Martin^31^, R. Nyanhete^3^, R. Wright^2^, R. Martin^62^, R. Davidson^34^, R. Cleaver^45^, S. Talukdar^16^, S. Butler^64^, J. Sampson^19^, S. Ribeiro^49^, S. Dell^46^, S. Mackenzie^32^, S. Hegarty^62^, S. Albaba^5^, S. McKee^36^, S. Palmer-Smith^19^, S. Heggarty^62^, S. MacParland^62^, S. Greville-Heygate^49^, S. Daniels^4^,S. Prapa^18^,S. Abbs^4^, S. Tennant^33^, S. Hardy^43^, S. MacMahon^49^, T. McVeigh^49^, T. Foo^49^, T. Bedenham^42^, T. Cranston^42,^ T. McDevitt^40^, V. Clowes^29^, V. Tripathi^23^, V. McConnell^62^, N. Woodwaer^45^, Y. Wallis^64^, Z. Kemp^49^, G. Mullan^62^, L. Pierson^62^, L. Rainey^62^, C. Joyce^59^, A. Timbs^41^, A-M. Reuther^3^, B. Frugtniet^47^, B. DeSouza^25^, C. Husher^3^, C. Lawn^22^, C. Corbett^63^, D. Nocera-Jijon^16^, D. Reay^31^, E. Cross^3^, F. Ryan^3^, H. Lindsay^6^, J. Oliver^6^, J. Dring^63^, J. Spiers^65^, J. Harper^23^, K. Ciucias^34^, L. Connolly^60^, M. Tsang^62^, R. Brown^6^, S. Shepherd^32^, S. Begum^16^, S. Daniels^3^, T. Tadiso^16^, T. Linton-Willoughby^4^, H. Heppell^35^, K. Sahan^61^, L. Worrillow^6^, Z. Allen^22^, C. Watt^34^,M. Hegarty^62^, R. Mitchell^6^, R. Coles^66^, G. Nickless^23^, E. Cojocaru^49^, I. Doal^64^, F. Sava^64^, C. McCarthy^62^, R. Jeeneea^63^, D. Goudie^20^, M. McConachie^20^, S. Botosneanu^5^, G. Kavanaugh^1^, K. Russell^10^, C. Sherlaw^63^, O. Tsoulaki^46^, C. Forde^5^, E. Petley^63^, A-B. Jones^1^, K. Oprych^16^, S. Pryde^67^, Z. Hyder^5^, N. Elkhateeb^18^, R. Braham^21^, L. Hanington^41^, C. Huntley^1^, R. Irving^51^, A. Sadan^23^, M. Ramos^22^, C. Elliot^35^, D. Wren^22^, D.Lobo^35^, J. McLean^68^, D. May^18^, L. Kearney^48^, T. Campbell^38^, K. Asakura^68^, L. Alwadi^19^, R. O’Shea^48^, J. Gabriel^42^, L. Chiecchio^3^, P. Bowman^44^, L.A. Sutton^48^, C. Walsh^23^, V. Cloke^69^, D. Ucanok^37^, J. Davies^65^, B. Pleasance^65^, E. Maguire^6^, A. Whaite^70^, S. Best^71^, S. Westbury^72^, A. Logan^62^, D. Navarajasegaran^71^, A. Bench^35^, P. Wightman^34^, A. Cartwright^2^, E. Higgs^41^, J.Bott^42^, H. Whitehouse^5^, J. Stevens^58^, D. Martin^37^, J. Dunlop^68^, S. Thomas^73^, C. Sau^68^, S. Farndon^74^, N. Coleman^48^, P. Angelini^49^, M. Duff^59^, H. Massey^35^, C. Rowlands^1^, C. Garcia-Petit^68^, K. Gillespie^68^, A. Alder^68^, E. Middleton^68^, C. Cassidy^75^, N. Orfali^48^, A. Webb^3^, A. Luharia^64^, N. Walker^33^, J. Charlton^71^, A. Andreou^47^, J. Peddie^68^, M. Khan^23^, L. Wilkinson^31^, H. Bezuidenhout^47^, M. Edis^18^, A. Callard^29^, P. Ostrowski^76^, P. Moverley^51^, K. Bean^73^, A. Dunne^48^, A. Moleirinho^23^, S. Waller^5^, K. Cox^47^, L. Greensmith^28^, A. Brittle^5^, N. Gossan^5^, L. Freestone^4^, C. Shak^77^, T. Langford^75^, Y. Clinch^21^, H. Livesey^51^, S. Borland^46^, A. Joshi^47^, K. Wall^77^, A. Whitworth^46^, A.Wilsdon^37^, K. Edgerley^72^, S. Pugh^5^, N. Chrysochoidi^3^, S. Mutch^38^, C. McMullan^5^, Y. Johnston^78^, M. Muraru^77^, A. May^77^, R. Begum^77^, C. Smith^44^, R. Patel^47^, I. Bhatnagar^79^, A. Taylor^62^, D. Brown^69^, J. Willan^46^, S. Taylor^48^, K. Jones^21^, K. Cox^21^, C. Ramsden^75^, O. Taiwo^49^, J. Jaudzemaite^47^, R. Sharmin^47^, L. Young^34^, C.O’Dubhshlaine^35^, L. McSorley^80^, S. Lillis^23^, P. Alexopoulos^23^, E. Mortensson^65^, L. Kingham^4^, R. Moore^18^, M. Kosicka-Slawinska^81^, S. Aslam^65^, R. Wells^82^, A. Carter^82^, H. Warren^6^, E. Rolf^49^, H. Reed^75^, L. Pearce^83^, D. Lock^6^, F. Ali^66^, A. Kolozi^84^, N. White^48^, D. Wood^34^, C. Hayden^25^, W. Cheah^58^, J. Sims^6^, R. Heron^46^, J. Sibbring^4^, L. Elmhirst^35^, L. Mavrogiannis^6^, K. Oakhill^4^, L. Wang^28^, A. Singh^47^, K. Doal^21^, L. Kettle^63^, R. Salmon^63^, G. Thodi^48^, C. O’Brien^48^, C. Wragg^85^, N. Mannion^34^, S. Chu^63^, M. Ukash^75^, V. Steventon-Jones^46^, J. Fairley^84^, H. Northen^18^, D. Babu^75^, L. Donaghy^34^, J. Jimmy^48^, B. Matharu^18^, J. Beasley^42^, S. Waller^75^, C. Batterton^63^, G. Baker^4^, J. Trotman^4^, L. Jackson^11^, A. Visavadia^63^, M. Domeradzka^49^, M. Slater^22^, K. Annesley^18^, C. Andrews^1^, J. Doughty^18^, E. Wall^63^, S. Morosini^46^, E. Hanney^60^

^1^ Division of Genetics and Epidemiology, Institute of Cancer Research, Sutton, UK

^2^ Sheffield Diagnostic Genetics Service, NEY Genomic Laboratory Hub, Sheffield Children's NHS Foundation Trust, Sheffield, UK

^3^ Wessex Genetics Laboratory Service, University Hospital Southampton NHS Foundation Trust, Salisbury, UK

^4^ East Genomic Laboratory Hub, Cambridge University Hospitals Genomic Laboratory, Cambridge University NHS Foundation Trust, Cambridge, UK

^5^ Manchester Centre for Genomic Medicine and NW Laboratory Genetics Hub, Manchester University Hospitals NHS Foundation Trust, Manchester, UK

^6^ The Leeds Genetics Laboratory, NEY Genomic Laboratory Hub, Leeds Teaching Hospitals NHS Trust, Leeds, UK

^7^ Cancer Sciences, Faculty of Medicine, University of Southampton, Southampton, UK

^8^ Human Genetics and Genomic Medicine, Faculty of Medicine, University of Southampton, Southampton, UK

^9^ Department of Molecular Genetics, Royal Devon and Exeter NHS Foundation Trust, Exeter, UK

^10^ Genomics England, London, UK

^11^ University of Exeter Medical School, Exeter, UK

^12^ Division of Evolution & Genomic Sciences, The University of Manchester

^13^ Department of Medical Genetics, National Institute for Health, Research Cambridge Biomedical Research Centre, University of Cambridge, Cambridge, UK

^14^ Wessex Clinical Genetics Service, University Hospital Southampton NHS Foundation Trust, Southampton, UK

^15^ Clinical Ethics and Law Unit, University of Southampton, Southampton, UK

^16^ Department of Clinical Genetics, St. George's University Hospitals NHS Foundation Trust, London, UK

^17^ Public Health and Primary Care, Clinical Medicine, University of Cambridge, Cambridge, UK

^18^ Cambridge University Hospitals NHS Foundation Trust, Cambridge, UK

^19^ Institute of Medical Genetics, University Hospital of Wales, Cardiff and Vale University Health Board, Cardiff, UK

^20^ East of Scotland Regional Genetics Service, Level 6, Ninewells Hospital, Dundee

^21^ Great Ormond Street Hospital for Children NHS Foundation Trust, London, UK

^22^ North Thames Genomic Laboratory Hub, Great Ormond Street Hospital for Children NHS Foundation Trust, London, UK

^23^ Department of Clinical Genetics, Guy’s and St Thomas’ NHS Foundation Trust, London, UK

^24^ South East Genomic Laboratory Hub, Guy’s and St Thomas’ NHS Foundation Trust, London, UK

^25^ Genomic Medicine Service, Imperial College Healthcare NHS Trust, London, UK

^26^ Faculty of Medicine, Department of Surgery & Cancer, Imperial College London, London, UK

^27^ Institute of Neurology, UCL Queen Square Institute of Neurology, London, UK

^28^ Liverpool Women’s NHS Foundation Trust, Liverpool, UK

^29^ London North West University Healthcare NHS Trust, London, UK

^30^ Division of Evolution and Genomic Sciences, School of Biological Sciences, Faculty of Biology Medicine and Health, The University of Manchester, Manchester, UK.

^31^ The Newcastle upon Tyne Hospitals NHS Foundation Trust, Newcastle upon Tyne, UK

^32^ North East and Yorkshire Genomic Laboratory Hub, The Newcastle upon Tyne Hospitals NHS Foundation Trust, Newcastle upon Tyne, UK

^33^ NHS Grampian, Aberdeen, UK

^34^ NHS Greater Glasgow and Clyde, Glasgow, UK

^35^ NHS Lothian, Edinburgh, UK

^36^ Northern Ireland Regional Genetics Service, Belfast Health & Social Care Trust, Belfast, UK

^37^ Nottingham University Hospitals NHS Trust, Nottingham, UK

^38^ East Midlands and East of England Genomics Laboratory, Nottingham University Hospitals NHS Trust, Nottingham, UK

^39^ University of Otago, Otago, New Zealand

^40^ Our Lady's Children's Hospital, Crumlin, Dublin, Ireland

^41^ Clinical Genetics, Oxford University Hospitals NHS Foundation Trust, Oxford, UK

^42^ West Midlands, Oxford and Wessex Genomic Laboratory Hub, Oxford University Hospitals NHS Foundation Trust, Oxford, UK

^43^ Public Health England, London, UK

^44^ Royal Devon and Exeter NHS Foundation Trust, Exeter, UK

^45^ Royal Free London NHS Foundation Trust, London, UK

^46^ Sheffield Children's NHS Foundation Trust, Sheffield, UK

^47^ St George’s University Hospitals NHS Foundation Trust, London, UK

^48^ St James’s Hospital, Dublin, Ireland

^49^ Cancer Genetics Unit, The Royal Marsden NHS Foundation Trust, Sutton, London, UK

^50^ Trinity College Dublin, The University of Dublin, Ireland

^51^ University Hospital of Wales, Cardiff and Vale University Health Board, Cardiff, UK

^52^ University Hospitals Bristol NHS Foundation Trust, Bristol, UK

^53^ University Hospitals of Leicester NHS Trust, Leicester, UK

^54^ University Hospitals of Plymouth NHS Trust, Plymouth, UK

^55^ University of Manchester, Manchester, UK

^56^ Wessex Clinical Genetics Service, Princess Anne Hospital, Southampton, UK

^57^ Faculty of Medicine, University of Southampton, Southampton, UK

^58^ University Hospital Southampton NHS Foundation Trust, Southampton, UK

^59^ Cork University Hospital, Cork, Ireland

^60^ Children’s Health Ireland (CHI), Crumlin, Dublin, Ireland

^61^ The Ethox Centre, Oxford, UK

^62^ Belfast Health & Social Care Trust, Belfast, UK

^63^ Birmingham Women’s and Children’s NHS Foundation Trust, Birmingham, UK

^64^ Central and South Genomic Laboratory Hub, Birmingham Women’s and Children’s NHS Foundation Trust, Birmingham, UK

^65^ Bristol Genetics Laboratory, Pathology Sciences, Southmead Hospital, North Bristol NHS Trust, Bristol, United Kingdom

^66^ Northwick Park Hospital, Watford Rd, Harrow, UK

^67^ Chapel Allerton Hospital, Chapeltown Rd, Leeds, UK

^68^ NHS Tayside, UK

^69^ South East Scotland Genetic Service, Western General Hospital, Edinburgh, UK

^70^ Liverpool Centre for Genomic Medicine, Liverpool Women’s NHS Foundation Trust, Liverpool, UK

^71^ King’s College Hospital, London, UK

^72^ University Hospital Bristol and Weston NHS Foundation Trust, Bristol, UK

^73^ Sheffield Teaching Hospital NHS Foundation Trust, Sheffield, UK

^74^ Bristol Royal Hospital for Children, Bristol. UK

^75^ Manchester University Foundation Trust, Manchester, UK

^76^ North East Thames Clinical Genetics Service, London, UK

^77^ West Midlands Regional Genetics Laboratory, Birmingham Women’s Hospital, Birmingham, UK

^78^West of Scotland Centre for Genomic Medicine, Queen Elizabeth University Hospital, Glasgow, UK

^79^Oxford Centre for Genomic Medicine, Oxford University Hospitals NHS Foundation Trust, Oxford, UK

^80^St Vincent’s Hospital Group, Elm Park, Dublin, Ireland

^81^North West Thames Regional Genetics Service, St. Mark's Hospital, Harrow, UK

^82^Royal Liverpool University Hospital Trust, Liverpool, UK

^83^All Wales Medical Genetics Service, Cardiff, Wales, UK

^84^GenQA

^85^South West Genomic Laboratory Hub, Bristol Genetics Laboratory, Southmead Hospital, Bristol, UK
